# Retinal vascularization rate predicts retinopathy of prematurity and remains unaffected by low-dose bevacizumab treatment

**DOI:** 10.1101/2025.01.07.24318722

**Authors:** Emer Chang, Amandeep Josan, Ravi Purohit, Sher A Aslam, Caroline Hartley, Chetan K Patel, Kanmin Xue

**Affiliations:** Oxford Eye Hospital, Oxford University Hospitals NHS Foundation Trust, Oxford, UK; Nuffield Laboratory of Ophthalmology, Nuffield Department of Clinical Neurosciences, University of Oxford, Oxford, UK; Department of Paediatrics, University of Oxford, Oxford, UK; Ophthalmology Department, Great Ormond Street Hospital for Children NHS Foundation Trust, London, UK

**Keywords:** retinopathy of prematurity, ROP, ultra-widefield imaging, retinal vascularisation rate, machine learning

## Abstract

**Purpose:** To assess the rate of retinal vascularisation derived from ultra-widefield (UWF) imaging-based retinopathy of prematurity (ROP) screening as predictor of type 1 ROP and characterise the effect of anti-VEGF therapy on vascularisation rate.

**Design:** Retrospective, consecutive cohort study.

**Subjects:** 132 eyes of 76 premature infants with mean gestational age (GA) of 26.0(±2.0SD) weeks and birth weight (BW) of 815(±264) g, who underwent longitudinal UWF imaging for ROP screening.

**Setting/Venue:** Level 3 neonatal unit in Oxford, United Kingdom

**Methods:** The extent of retinal vascularisation on each UWF image was measured as the ratio between ‘disc-to-temporal vascular front’ and ‘disc-to-fovea’ distance along a straight line bisecting the vascular arcades. Measurements from ≥3 timepoints plotted against post-menstrual age (PMA) enabled calculation of temporal vascularisation rate (TVR) for each eye. Using TVR, GA and BW as predictors, a machine learning model was created to classify eyes as either Group AB (no ROP and type 2 ROP) or Group C (type 1 ROP). The model was validated in a withheld cohort of 32 eyes (19 infants) of which 8 eyes (5 infants) required treatment. TVR in 37 eyes (20 infants) was compared before and after ultra-low-dose (0.16 mg) intravitreal bevacizumab treatment.

**Main Outcome Measures:** Rate of retinal vascularisation.

**Results:** Slower retinal vascularisation correlated with increasing ROP severity, with TVR being 29% slower in Group C eyes (n=50) than Group AB eyes (n=33 no ROP and n=49 type 2 ROP) (p=0.04). Our model correctly predicted ROP outcomes of 30/32 eyes, achieving a balanced accuracy of 95.8%. No significant change in TVR was found before and after bevacizumab treatment with mean post-treatment imaging follow-up of 7.7(±7.9) weeks (p=0.60 right eyes, p=0.71 left eyes).

**Conclusions:** UWF imaging-based ROP screening enables quantification of retinal vascularisation rate, which can provide early prediction of type 1 ROP independent of BW and GA. Rate of physiological retinal vascularisation does not appear to be significantly affected by ultra-low-dose anti-VEGF treatment, which has significant implications for the development of peripheral avascular retina and timing of anti-VEGF intervention to prevent disease progression in high risk infants.

## INTRODUCTION

Retinopathy of prematurity (ROP) is a leading cause of childhood blindness worldwide (Solebo 2017). It is characterised by aberrant retinal vascular development that progresses through two main phases: (i) delayed retinal vascularisation in phase 1 results in peripheral avascular retina (PAR), which stimulates release of angiogenic factors; followed by (ii) a vasoproliferative phase 2 associated with extraretinal neovascularisation and subsequent tractional retinal detachment (Patz et al 1952, Ashton 1954, Hartnett and Penn 2012). ROP causes visual impairment in over 30 000 preterm babies globally (Blencowe et al. 2013) with rising incidence over the past decades owing to improved survival of extreme preterm infants (e.g. those born as early as 22 weeks) (Bhatnagar et al. 2023).

Timely detection of ROP enables treatment by anti-vascular endothelial growth factor (anti-VEGF) therapy or laser photocoagulation to prevent sight loss (“UK screening of ROP guideline” 2022). Low gestational age (GA <31 weeks) and birthweight (BW ≤1500g) are major risk factors for developing ROP, thus constitute the inclusion criteria for ROP screening (“UK screening of ROP guideline” 2022). Other risk factors include oxygen supplementation, slow postnatal weight gain, sepsis, and respiratory distress (Dammann, Hartnett, and Stahl 2023). While many of these factors are interlinked, their individual predictive power for ROP occurrence remains poorly defined.

The current ROP grading system (based primarily on Zones I-III, stages 0-5 and normal to plus disease spectrum) (Chiang et al 2021) is applied during ROP screening examinations using either binocular indirect ophthalmoscopy (BIO) or digital retinal imaging. BIO is a technically demanding skill in premature infants and is mainly focused on detecting features of vasoproliferation, such as stage 3 (elevated neovascular ridge) and plus disease (vessel dilation and tortuosity), which pertains to the ROP treatment threshold.

However, indirect ophthalmoscopy is less suited to assessing delayed retinal vascularisation, which can only be gleaned approximately from 33-34 weeks post-menstrual age (PMA) at which vessels reach the edge of Zone I or Zone II (Jang and Kim 2020).

Wide-field retinal imaging could enable accurate assessment and longitudinal monitoring of the rate of retinal vascularisation from birth. Contact retinal imaging such as the 130⁰ RetCam 3 Wide-field Digital Imaging System (Natus Medical Inc., Pleasanton, CA, USA) provides limited ability to visualise the posterior pole and far peripheral retina in a single imaging field (Kumar et al. 2021; Padhi et al. 2022; Solans Pérez De Larraya et al. 2019; Solans Pérez de Larraya et al. 2018). Non-contact 200-degree ultra-widefield (UWF) retinal imaging (e.g. Optomap, Optos plc., Dunfermline, Scotland, UK) could potentially overcome this limitation to enable consistent and accurate quantification of the rate of retinal vascularisation over time. We have previously demonstrated the utility of UWF imaging for ROP assessment with the infants being held to the camera using the ‘flying baby’ technique (Patel et al. 2013). We have more recently transitioned from BIO examination to UWF imaging as the default method of ROP screening at our level 3 neonatal unit using an Optos California imaging system mounted on a mobile trolley with onboard battery.

In this study, we aimed to devise a practical method for measuring the rate of retinal vascularisation in a retrospective analysis of serial UWF images obtained from routine ROP screening. Using retinal vascularisation rate, GA and BW as predictors, we aimed to create a machine learning based gradient boosting model for predicting ROP requiring treatment (type 1 ROP) which demonstrates high accuracy in a separate validation dataset.

The presence of periphery avascular retina (PAR) has been reported in a high proportion (91-100%) of infants who have received intravitreal anti-VEGF therapy for ROP (Toy et al 2016; Lepore et al 2018). However, the causal relationship between anti-VEGF treatment and PAR remains unclear since PAR may be associated with ROP itself. Nonetheless, in clinical practice, concerns about potential vascular arrest following anti-VEGF treatment may lead clinicians to hold off anti-VEGF intervention in borderline treatment-requiring cases with posterior disease, e.g. posterior Zone II stage 2 with Plus (UK ROP guideline: ‘Treating Retinopathy of Prematurity in the UK 2022’). As part of this study, we aimed to quantify and compare the rate of retinal vascularisation before and after intravitreal anti-VEGF treatment for type 1 ROP.

## METHODS

### Data collection and study eligibility criteria

The ROP database at the Oxford University Hospitals NHS Foundation Trust (a Level 3 Neonatal Care Unit, John Radcliffe Hospital, Oxford, UK) was used to retrospectively identify all infants who underwent imaging-based ROP screening in accordance with UK guidelines (“Treating Retinopathy of Prematurity in the UK” 2022) between May 2019 to February 2024. Ethics approval and consent to use patient data for analyses was approved by the Oxford University Hospitals Integrated Governance System (Oxford University Hospitals clinical audit approval no. 9080) and were conducted as an internal retrospective clinical audit. Due to the retrospective and anonymous nature of this audit, informed consent was waived by the ethics committee. Inclusion criteria for the study included availability of at least three 200-degree ultra-widefield (UWF) retinal images (Optos California) recorded at least one-week apart, with known outcomes of ROP screening that were classified into three categories:

- *Group A* (no ROP): eyes that were recorded as having full or Zone 3 retinal vascularisation without developing any ROP and discharged from ROP screening;
- *Group B* (ROP not requiring treatment): eyes that had complete regression of ROP without reaching treatment threshold (i.e. type 2 ROP);
- *Group C* (ROP requiring treatment): eyes with type 1 ROP that required treatment for ROP (either anti-VEGF therapy or laser).

Infants without definitive ROP outcomes (e.g. infants that were transferred to other units or passed away) and those with other significant ocular co-morbidity were excluded from the analysis. Eyes that did not have serial UWF imaging of sufficient quality to measure the extent of temporal retinal vascularisation were also excluded.

The dataset was used for the following two major analyses:

*Analysis 1* – Retinal vascularisation rates as a predictor of type 1 ROP. To develop our machine learning model for predicting type 1 ROP, we excluded retinal images obtained after any ROP treatment or after 40 weeks PMA to avoid bias, as fewer scans beyond 40 weeks PMA would be associated with infants who do not develop any ROP (Pruett, Ruland, and Donahue 2023). Infants that started ROP screening from May 2019 up to the end of March 2023 were used to train the ROP prediction model. Infants not included in model training, that started screening from April 2023 to February 2024, were used as an independent test set to validate the predictive model.

*Analysis 2* - Effects of anti-VEGF therapy on retinal vascularisation. To determine the effect of anti-VEGF treatment on the rate of retinal vascularisation, we included all eyes with type 1 ROP with at least two UWF images captured before and two images after intravitreal injection of bevacizumab (0.16 mg in 0.025 ml). For this analysis, UWF images captured after 40 weeks PMA were included in order to assess the long-term effects of anti-VEGF therapy on retinal vascularisation.

Alongside the UWF retinal images, the following information were also collected for each baby: gestational age (GA), birth weight (BW), ROP outcome based on International Classification of ROP(ICROP 2005), PMA at each ROP screening timepoint, and PMA at the time of anti-VEGF injection (if applicable).

### Ultra-widefield retinal image analysis

For each UWF retinal image, the advancement of temporal vascular front was measured as the ratio between the distance from ‘optic disc centre-to-temporal vascular front’ and ‘optic disc centre-to-fovea’ along a straight line that bisects the superior and inferior venous arcades. Identifying the fovea in preterm infants can be challenging as it is not as well-defined due to delayed or altered development in prematurity and ROP (Isenberg 1986, Akula et al 2020). For the purpose of consistent measurements across longitudinal UWF images, we defined the fovea as the point that bisects the widest distance between the superior and inferior venous arcades (Wilson et al 2006). Retinal veins were chosen instead of arteries, as the arteries tend to become more tortuous in pre-plus or plus disease (ICROP 1984). The red-free filter was used to provide improved contrast between vessels and the surrounding tissue to aid identification of vascular front and venous arcades. To validate this approach, we performed fovea localisation using the same method on confocal scanning laser ophthalmoscopy (cSLO) images accompanying Heidelberg Flex optical coherence tomography (Flex OCT, Spectralis, Heidelberg Engineering, Heidelberg, Germany) in 13 eyes of 10 premature infants (**Fig.1**). The location of ‘presumed’ fovea on the cSLO image was compared with the ‘actual’ anatomical fovea as identified on the corresponding OCT. This showed a mean discrepancy of only 0.4 mm (SD=0.36) between the presumed and actual fovea locations (**Supplementary Table S1**), thus validating our method for consistently identifying the fovea on UWF images.

**Fig.1.**
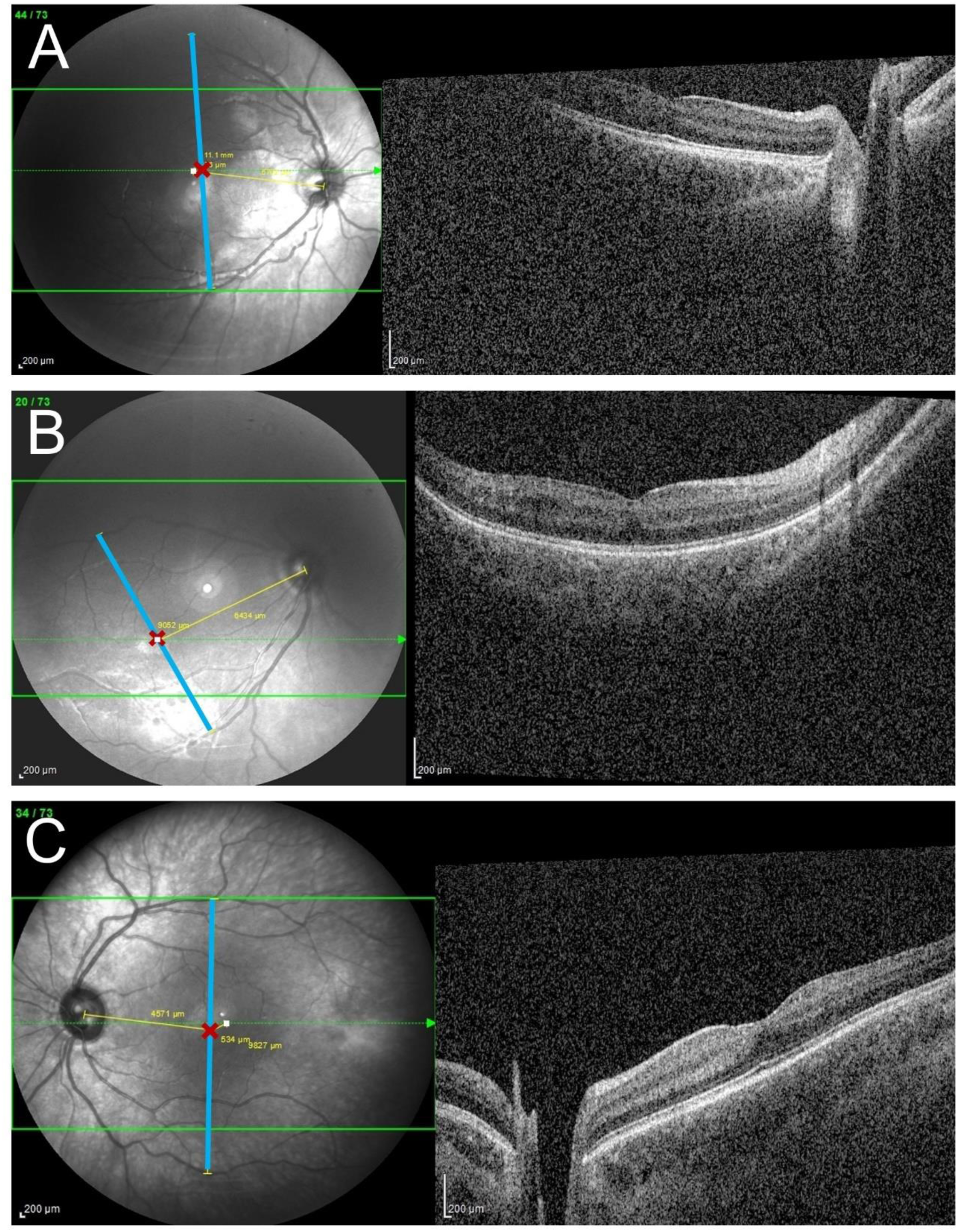
Validation of method for fovea localisation in premature infants using Flex OCT. ‘Presumed’ fovea (red cross) was defined on the Spectralis scanning laser ophthalmoscopy (SLO) images as the point that bisects the widest distance between the superior and inferior venous arcades (intersection between the yellow and blue lines). ‘Actual’ (anatomical) fovea (white dot) was identified on the corresponding Spectralis Flex OCT and transferred to the SLO images using Heidelberg Eye Explorer (HEYEX) marker tool. A mean discrepancy of 0.40 mm (SD=0.36) was achieved across 13 eyes of 10 premature infants imaged. Three representative examples are shown: (**A**) right eye of a baby born at 26-27 weeks with a discrepancy of 0.30 mm between presumed and actual fovea locations; (**B**) right eye of a baby born at 24-25 weeks with 0 mm discrepancy; (**C**) left eye of a baby born at 24-25 weeks with 0.53 mm discrepancy. To maintain anonymity, gestational age is presented as age range in weeks.

The disc-to-fovea distance (D-F) was chosen as the reference unit of distance measurement so that subsequent disc-to-temporal vascular front distance measurements could be expressed as a ratio to D-F. This is consistent with the methodology used in previous studies (Lorenz et al 2017, Zhang et al 2024). In addition, De Silva et al demonstrated that longitudinal D-F measurements did not change significantly over 9 weeks in preterm and full-term infants (De Silva et al 2006). This normalisation helps to eliminate the effects of small changes in image magnification/angle and axial length growth on calliper measurements taken from images of the same eye at different time points (**Supplementary Fig.S1A**). Using this method, temporal vascularisation was observed to progress in a linear fashion in untreated eyes of premature infants (**Supplementary Fig.S1B**). It should be noted that while all eyes grow in size between 32-52 weeks gestation (approx. 0.15mm/week, as described by Cook et al. 2008), the difference in the rate of axial length increase between normal and stage 1-3 ROP eyes is minimal and, importantly all eyes follow linear growth trajectories over the period in question. The other practical benefit of using D-F distance as the reference unit is to allow inference of vascularisation to the edge of Zone 1, since the diameter of Zone 1 is defined as twice the distance from fovea to disc centre (i.e. 2x D-F) (**Supplementary Fig.S1B**). If a vascular ridge was present, the location of the vascular front was defined as the centre (or peak) of the ridge. All distance measurements were made using the calliper tool within OptosAdvance (Optos plc.) with in-built peripheral retinal curvature correction (Sagong et al. 2015; Nagiel et al. 2016; Tan et al. 2016). These measurements were reviewed twice by investigator EC and reviewed by a second independent investigator (RP) to assess inter-rater correlation and achieve a consensus. Measurements of the disc-to-temporal vascular front were regressed against PMA at the time of imaging and the gradient of the line of best fit taken as the temporal vascularisation rate (TVR) for an individual eye.

### Statistical analysis

Statistical analysis was conducted using the programming software R (v4.2.1). Linear mixed effect modelling was used to account for repeated measures using the ‘lme4’ package (Bates et al. 2015). Model performance and diagnostics were carried out using the ‘performance’ package (Lüdecke et al. 2021) and effect size analysis for the linear mixed effects model was performed using the ‘simr’ package (Green and Macleod 2016).

Temporal vascularisation rate (TVR) for each eye was calculated using a linear regression slope analysis of the advancement of temporal vascular front versus PMA at imaging. As part of our preliminary work, the disc-to-nasal vascular front was also measured in a similar manner to disc-to-temporal vascular front in eyes that had UWF images that captured nasal peripheral retina on more than one occasion (using D-F distance as the common unit). The advancement of nasal vascular front was found to correlate strongly with temporal vascular front with a correlation coefficient of 0.91 (**Supplementary Fig.S2**). However, there was insufficient longitudinal data to calculate the nasal vascularisation rate (NVR) for most eyes due to technical challenge of consistently imaging the peripheral nasal retina. Therefore, all subsequent analyses utilised TVR as a surrogate measure of the overall rate of retinal vascularisation for each eye.

To compare vascular growth rates between eyes that did not require treatment and those that did require treatment, Groups A and B eyes were combined to represent all those with no ROP or type 2 ROP (not requiring treatment). Group C included infants with type 1 ROP for which treatment is recommended as per ICROP guidelines (Chiang et al. 2021).

To detect differences in the rate of temporal retinal vascularisation as a surrogate marker of overall vascularisation rates (slopes) between these two groups, a linear mixed effect model with random slopes and random intercepts was fitted using both eyes and patient ID as the nesting variables to account for similarities between right and left eyes in each patient and the repeated measures nature of the longitudinal data. The fixed effect independent predictor variables investigated were PMA, GA, BW and ROP group. The R package ‘glmultì was used to compare all possible combinations of predictors along with their interactions (Calcagno 2020). Model comparisons were performed and evaluated using Akaike information criterion (AIC) and Bayes information criterion (BIC) to arrive at the best model to describe the independent variable of temporal vessel extent. The linear mixed effects analysis omnibus test was followed by Tukey post-hoc comparison of slopes. Model assumptions were verified using visual normal distribution checks, q-q plot analysis, symmetry of histograms and assessment of heteroscedasticity (**Supplementary Fig.S3**). Where model assumptions were violated, such as in the comparison of TVR pre and post anti-VEGF treatment, robust linear mixed modelling was used. Robust models mitigate the effects of the outliers by applying a trimmed weighting to extreme outlier patients. An alternative Bayesian framework was also investigated; however the robust model fit was found to be superior, likely as a result of the presence of extreme outliers and the setting of vague priors due to a lack of existing knowledge in this novel domain.

Monte Carlo simulation adjusting effect sizes was implemented to determine the minimum effect size that could be reasonably detected at an alpha level of 0.05 and 90% power when compared to a null model.

### Development of ROP classification prediction model

Gradient boosting is an ensemble machine learning technique which generates a large number of decision trees sequentially, each learning from the last, in order to accurately classify according to the observed predictors. We developed a model using gradient boosting machine learning with the predictors, BW, GA and TVR (slope coefficient of the temporal vascularisation extent versus postmenstrual age), to predict whether a baby belongs to one of two groups: Group AB (no treatment) or Group C (ROP requiring treatment). We compared three models for prediction: one with TVR alone, one with BW and GA, and one model with all three predictors.

We chose to use a gradient boosting rather than random forest model due to the presence of nested data - the right and left eyes of each patient were included in the classification model. We used the package ‘GPBoost’ which combines linear mixed effects modelling and tree boosting to train the fixed effects and assign patient ID as the random effect to account for nested eyes (Sigrist 2021; Sigrist et al. 2024). GPBoost uses cross validation to tune the hyperparameters on the training dataset. Precision-recall curve (PRC) with area under the PRC (AUPRC) were calculated in addition to the receiver operating curves (ROC) and AUROC to assess each model from the training set. Both ROC and PR curves use sensitivity but the second axis differs: ROC uses false positive rate (1-specificity) whereas PRC uses precision defined as how many true positives out of all that have been predicted as positives. Whilst AUROC is more familiar to most, the AUPRC is a more suitable measure where there is class imbalance, particularly where there are far more negative cases than positive (Davis and Goadrich 2006, Saito 2015). The moderate imbalance between non-treatment group (n=48) and ROP requiring treatment group (n=28) in our dataset justifies the use of AUPRC over AUROC.

Bernoulli probit likelihood distribution was used for this binary classification training model and the optimizer employed was Nesterov-accelerated gradient descent. The gradient boosting predictive model was then tested using a fully independent validation set and assessed using balanced accuracy.

To establish whether TVR provides predictive information above that of BW and GA, we used a variable inflation factor analysis from the ‘car’ package (Fox and Weisberg 2019) that assessed variable importance and the presence of collinearity. This assesses multicollinearity between the predictors (GA, BW, TVR) by performing pairwise comparisons of every combination of predictors and analysing each correlation coefficient obtained as a result.

## RESULTS

### Demographics of patient groups

A total of 1204 infants underwent ROP screening during May 2019 to February 2024. Of these, 95 infants were included in the imaging analyses. 1109 infants were excluded mainly due to gradual transition from BIO to imaging based ROP screening, transfer to other units and insufficient number of UWF images.

Data from 76 infants (132 eyes) that started ROP screening from May 2019 to March 2023 were included in the training data set, and so used to investigate the rate of retinal vessel growth in infants with and without ROP requiring treatment and develop a predictive model. Note that the number of eyes to infants is not at 2:1 ratio due to exclusion of eyes with insufficient quality of UWF images for the delineation of temporal vascular front. Their mean GA was 26.0 (SD ±2.0) weeks and mean BW 815 (±264) g. Of the 76 infants, 22 infants (33 eyes) who did not develop any ROP were assigned to Group A; 28 infants (49 eyes) who had type 2 ROP were assigned to Group B; and 28 infants (50 eyes) with type 1 ROP requiring treatment were assigned to Group C. Representative UWF images for Groups A, B and C are shown in **Fig.2A**.

**Fig.2.**
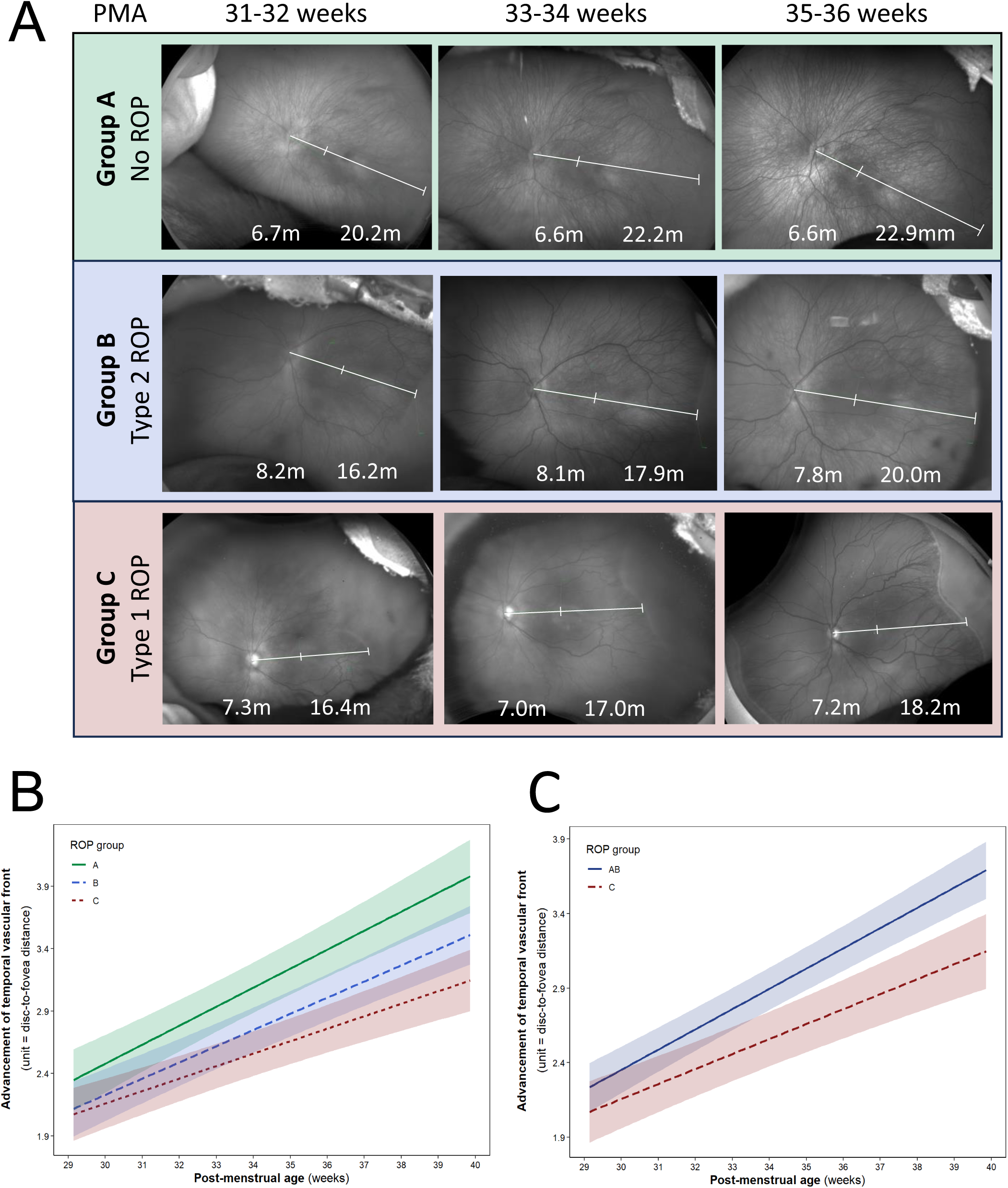
Correlation between temporal retinal vascularisation rate (TVR) and ROP occurrence. (**A**) Representative red-free UWF image series from consecutive ROP screening visits of eyes categorised as Group A (no ROP at any timepoint), Group B (type 2 ROP) and Group C (type 1 ROP). For each image, measurements (mm) of the distance from ‘disc centre to fovea’ (x) and ‘disc centre to temporal vascular front’ (y) are taken along a straight line that bisects the widest distance between the superior and inferior venous arcades. This enables expression of the extent of advancement of temporal vascularisation at each timepoint as a ratio (x/y). (**B**) Comparison of advancement in temporal vascular front relative to postmenstrual age (weeks) between Groups A (green), Group B (blue) and Group C (red). The slope represents the temporal vascularisation rate (TVR), which serves as a surrogate marker for the rate of retinal vascularisation for each group. Error bars represent 95% CI calculated at the ‘within subject’ level using the linear mixed model. (**C**) Comparison of advancement in temporal vascular front relative to postmenstrual age between the combined Group AB (no ROP treatment required, dark blue) and Group C (ROP requiring treatment, red). Mean TVR (slope) of Group AB eyes is significantly greater than Group C eyes (p=0.04).

Patients in Group A and Group B were subsequently merged into a combined Group AB, which represents all infants who did not require ROP treatment. Group AB consisted of 82 eyes from 48 infants. There is a discrepancy of 2 infants in the combined group, as these infants had one eye in Group A and the other eye in Group B.

The predictive model was tested using a fully independent test set containing data from 19 infants (32 eyes) that underwent ROP screening from April 2023 to February 2024 with all inclusion criteria fulfilled. Their mean GA was 26.4 (SD ±1.5) weeks and mean BW 803 (±240) g. Among these, 14 infants (24 eyes) did not require any treatment for ROP (Group AB) while 5 infants (8 eyes) required treated for ROP (Group C). The demographics of training and test data sets are summarised in **Table 1**.

**Table 1.** Demographics for infants in training and test groups. GA = gestational age; BW= birthweight.

|  | Training set (n=76 infants) | Test set (n=19 infants) |
| --- | --- | --- |
| Mean GA (±SD) (weeks) | 26.0 (2.0) | 26.4 (1.5) |
| Mean BW (±SD) (g) | 815 (264) | 803 (240) |
| No. of infants with type 1 ROP (%) | 28 (37) | 5 (26) |

#### Analysis 1

##### Slow rate of advancement of temporal retinal vascular front is associated with ROP

Across the training cohort, we found a trend for the TVR to decrease with increasing severity of ROP: the mean (±SEM) TVR for Group A eyes was 0.15 (±0.017) disc-to-fovea distance per week (D-F/week), Group B (ROP not requiring treatment) was 0.13 (±0.013) D-F/week, and Group C (ROP requiring treatment) 0.10 (±0.013) D-F/week (**Fig.2B**). Omnibus ANOVA test revealed significant differences in TVR (p=0.049), however, post-hoc testing with Tukey’s correction revealed no significant pairwise differences in TVR between Group A and B (p=0.60), between Group B and C (p=0.26), or between Group A and C (p=0.059). Given the clinical importance of distinguishing treatment requiring versus non-treatment requiring ROP and the somewhat subjective distinction between no ROP and stage 1 ROP, we merged Group A and B eyes into a combined Group AB (i.e. all eyes that did not require any therapeutic intervention) and compared this group with Group C (**Fig.2C**). In this case, the TVR of Group C was significantly slower (by 29%) than the TVR of Group AB (p=0.04, TVR of Group AB = 0.14±0.01 D-F/week, Group C = 0.10±0.014 D-F/week).

##### Vascularisation rate as a predictor of type 1 ROP

Given the significant difference in TVR between infants with and without ROP treatment, we hypothesised that TVR, calculated from at least two successive ROP screening visits, could be used as an independent predictor of type 1 ROP, over and above decisions based on GA and BW alone. A machine learning predictive model was developed using the training dataset, comparing three models incorporating (1) TVR only, (2) BW+GA, or (3) TVR+BW+GA as predictors (**Fig.3A**). The models were then tested using a fully independent test set as detailed above.

**Fig.3.**
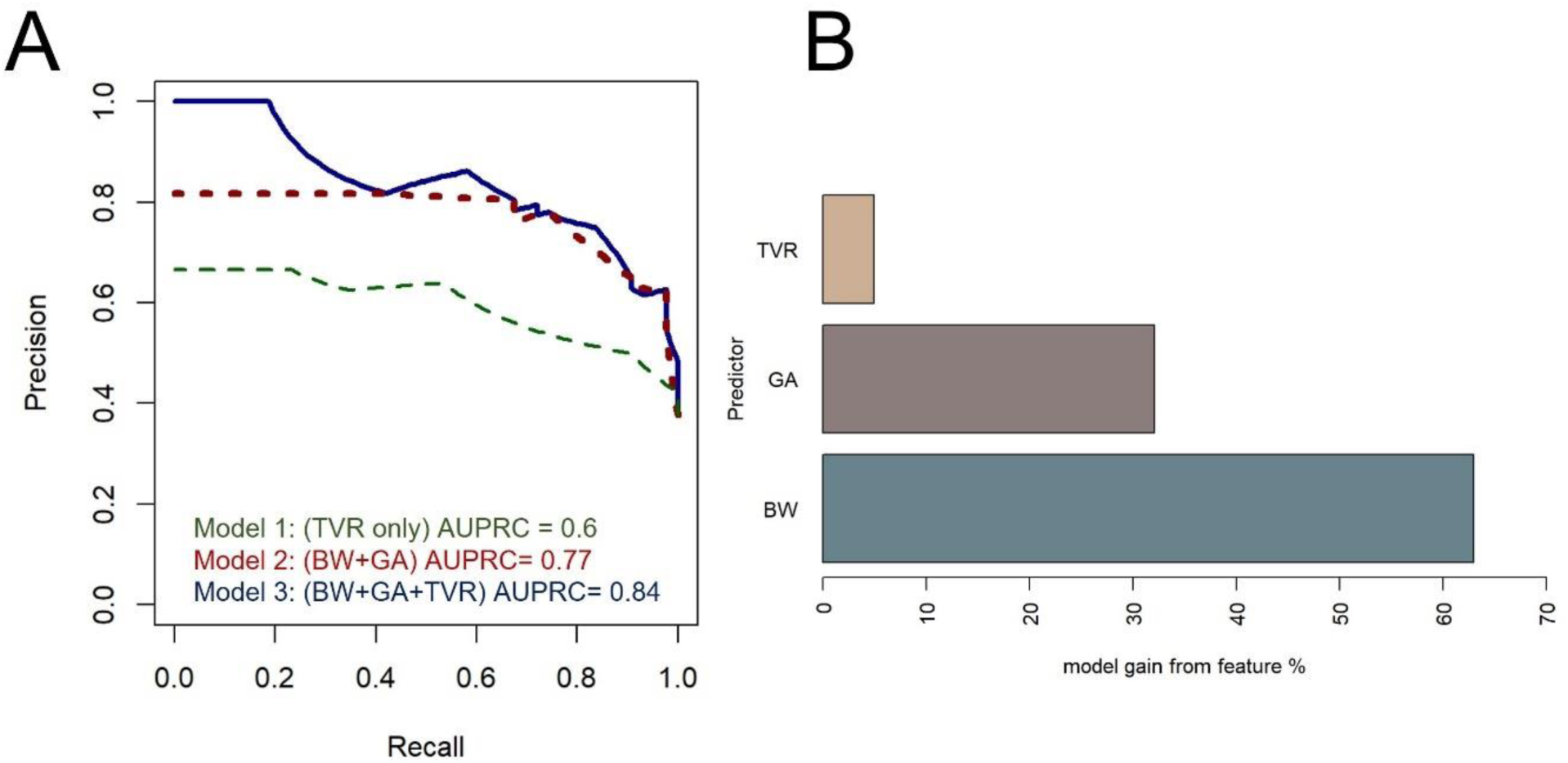
Temporal retinal vascularisation rate predicts Type 1 ROP. (**A**) Comparison of the precision-recall curves for 3 different prediction models. AUPRC: area under the precision-recall curve. Dark blue solid line represents model with predictors as birthweight (BW), gestational age (GA) and temporal vascularisation rate (TVR) that was used for further prediction. (**B**) Feature importance showing relative contributions of each predictor for the classification between treatment versus no treatment groups.

Whilst the combined predictive model (TVR+BW+GA) showed the best performance with an area under the precision recall curve (AUPRC) of 0.84 (**Fig.3A**) (AUROC 0.90, **Supplementary Fig.S4**), the model with TVR alone still performed well with an AUPRC 0.6 (AUROC 0.75). In the combined model, the model attributed percentage importance to each predictor for classifying type 1 ROP as 63% to BW, 32% to GA, and 5% to TVR (**Fig.3B**). Moreover, variable inflation factor analysis confirmed that TVR independently contributes to the predictive accuracy of the model with minimal collinearity with BW (r=0.16) or GA (r=0.10). This is in contrast to moderate/high collinearity between BW and GA (r=0.70).

In the independent test set, the combined model correctly predicted the outcomes of 30/32 eyes based on BW, GA and TVR (**Fig.3A** & **Table 2**). The balanced accuracy on the test data was 95.8%. The incorrect predictions were both eyes of patient Test15, which were predicted as belonging to Group C when the actual outcome was Group AB.

**Table 2.** Predicting type 1 ROP in a test dataset using a gradient boosting model.

| ID | Eye | GA (weeks) | BW (g) | TVR<br>(D-F/week) | Actual<br>ROP group | Model prediction<br>(TVR+GA+BW) |
| --- | --- | --- | --- | --- | --- | --- |
| Test1 | OS | 26-27 | 1,080 | 0.059 | AB | AB |
| Test1 | OD | 26-27 | 1,080 | 0.162 | AB | AB |
| Test2 | OS | 26-27 | 1,025 | 0.150 | AB | AB |
| Test2 | OD | 26-27 | 1,025 | 0.114 | AB | AB |
| Test3 | OD | 26-27 | 1,085 | 0.099 | AB | AB |
| Test4 | OS | 26-27 | 680 | 0.148 | AB | AB |
| Test4 | OD | 26-27 | 680 | 0.414 | AB | AB |
| Test5 | OS | 28-29 | 1,185 | 0.105 | AB | AB |
| Test5 | OD | 28-29 | 1,185 | 0.099 | AB | AB |
| Test6 | OD | 26-27 | 455 | 0.030 | C | C |
| Test7 | OS | 24-25 | 390 | 0.137 | C | C |
| Test7 | OD | 24-25 | 390 | 0.068 | C | C |
| Test8 | OS | 26-27 | 725 | 0.092 | AB | AB |
| Test8 | OD | 26-27 | 725 | 0.078 | AB | AB |
| Test9 | OD | 26-27 | 540 | 0.130 | AB | AB |
| Test10 | OS | 26-27 | 755 | 0.083 | C | C |
| Test10 | OD | 26-27 | 755 | 0.054 | C | C |
| Test11 | OS | 26-27 | 980 | 0.070 | AB | AB |
| Test11 | OD | 26-27 | 980 | 0.080 | AB | AB |
| Test12 | OS | 28-29 | 1,090 | 0.281 | AB | AB |
| Test12 | OD | 28-29 | 1,090 | 0.685 | AB | AB |
| Test13 | OD | 28-29 | 1,035 | 0.152 | AB | AB |
| Test14 | OD | 24-25 | 540 | 0.046 | C | C |
| Test15 | OS | 24-25 | 650 | 0.102 | AB | C |
| Test15 | OD | 24-25 | 650 | 0.131 | AB | C |
| Test16 | OS | 28-29 | 880 | 0.004 | AB | AB |
| Test16 | OD | 28-29 | 880 | 0.058 | AB | AB |
| Test17 | OS | 26-27 | 875 | 0.210 | AB | AB |
| Test17 | OD | 26-27 | 875 | 0.139 | AB | AB |
| Test18 | OS | 26-27 | 690 | 0.365 | AB | AB |
| Test19 | OS | 24-25 | 590 | 0.066 | C | C |
| Test19 | OD | 24-25 | 590 | 0.129 | C | C |

GA = gestational age, presented as age range in weeks to maintain anonymity; BW = birthweight. TVR = temporal vascularisation rate. D-F= disc-to-fovea distance. AB = no ROP + type 2 ROP (no treatment required group). C= type 1 ROP (treatment requiring ROP group). 'Test15' highlights a case of discrepancy between model prediction and actual ROP categorisation.

#### Analysis 2

##### Effects of anti-VEGF therapy on retinal vascularisation rate

To quantify the effect of anti-VEGF therapy on retinal vascularisation rate, we compared the TVR before and after treatment. Within our dataset, there were 37 eyes (from 20 infants) that received intravitreal bevacizumab injection (0.16 mg in 0.025 ml per eye) for ROP as well as having at least two UWF imaging timepoints before and two after the treatment (**Fig.4A**). Their mean (±SD) GA at birth was 24.2 (±0.9) weeks and BW 599 (±124) g. The mean follow-up length after anti-VEGF injection was 7.7 (±7.9) weeks; minimum follow-up 2 days and maximum follow-up 43 weeks.

**Fig.4.**
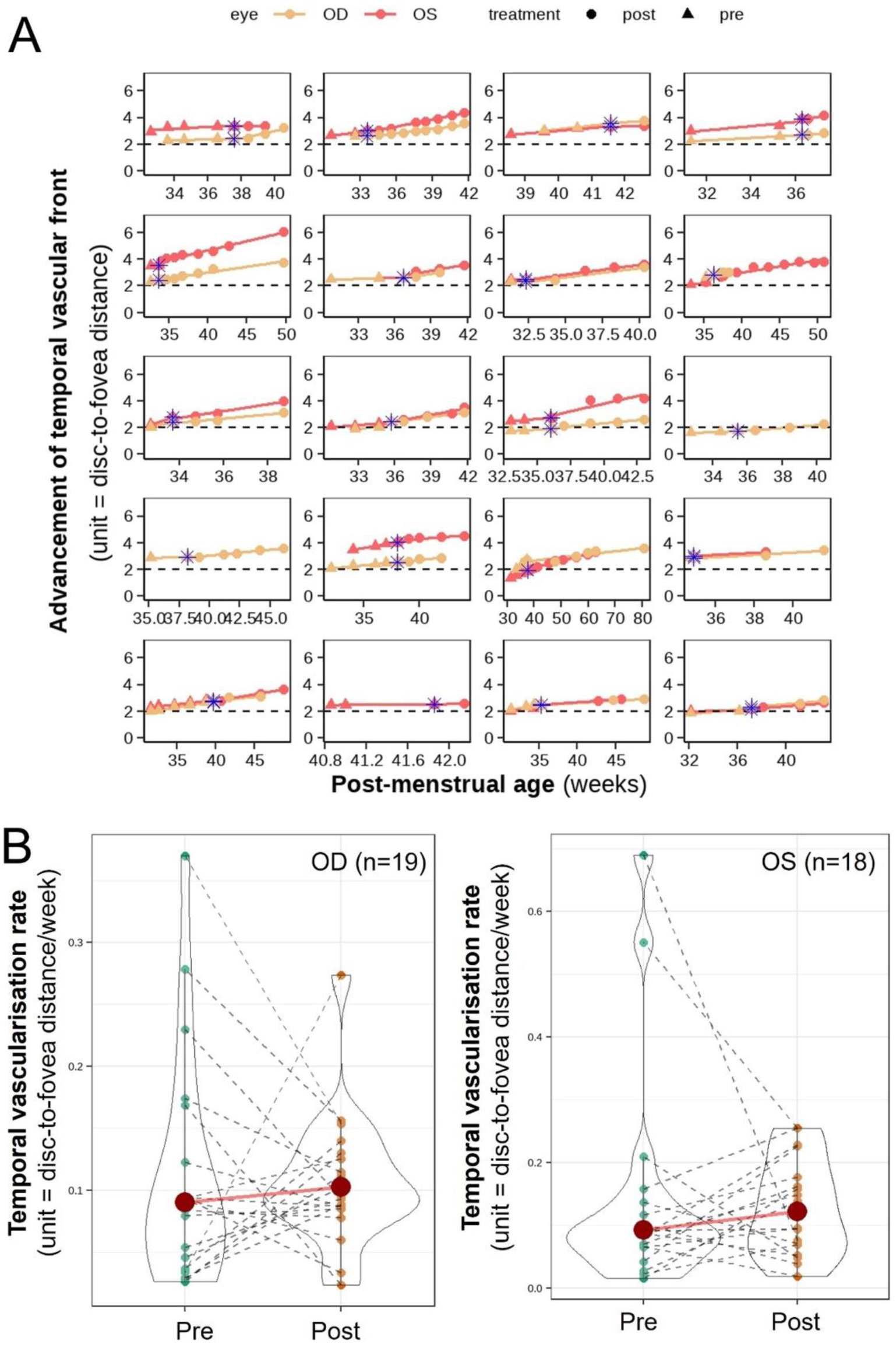
No significant change in temporal retinal vascularisation rate (TVR) before and after anti-VEGF therapy for ROP. (**A**) Comparison of advancement of temporal vascular front before and after bevacizumab (0.16 mg) injection in 20 infants (37 eyes). Each plot is from a different baby. Each triangle (pre-treatment) or circle (post-treatment) denotes an ultra-widefield (Optomap) imaging timepoint. Solid coloured line is the regression slope. Blue asterisk denotes the postmenstrual age at which anti-VEGF injection was given. Slope to the left of the asterisk is the TVR pre-treatment and slope to the right of the blue asterisk is the TVR post-treatment. Dotted line at y=2 where the ratio of disc-to-temporal vascular front distance to disc-to-fovea distance is 2, corresponds to the dividing line between Zone I and Zone II. (**B**) Violin plots comparing temporal vascularisation rate before and after bevacizumab (0.16 mg) injection for 19 right eyes and 18 left eyes. OD = right eye; OS = left eye. Each dotted line represents a single eye with temporal vascularisation before (green dot) and after treatment (orange dot). The red circle represents the mean temporal vascularisation rate (disc-to-fovea distance (D-F)/week).

Linear mixed model analysis showed no significant difference in TVR before and after anti-VEGF injection for both the right eyes (p=0.60) and the left eyes (p=0.71). Patients individually showed a mixture of changes (both increases and decreases) in TVR post-treatment, but collectively this was not statistically significant (**Fig.4B**). Post-hoc effect size analysis using Monte Carlo simulation demonstrated that there was sufficient sample size to detect a 3.3% (95% CI: 2.8-3.5) change in TVR after treatment (at alpha=0.05 and power=90%). Therefore, we conclude that anti-VEGF therapy does not significantly alter the rate of temporal retinal vascularisation.

## DISCUSSION

Given that delayed retinal vascularisation is a common end result of all major ROP risk factors, we hypothesised that its direct quantification through longitudinal UWF imaging could enable prediction of type 1 ROP. Our results demonstrate that slow rate of temporal vascularisation is correlated with ROP severity and can be used, along with BW and GA, to predict eyes that develop type 1 ROP with 96% accuracy. The results demonstrate the benefit of using retinal vascularisation rate derived from UWF imaging-based ROP screening, highlighting its ability to provide novel quantitative insights into disease progression and treatment response early in the screening/monitoring process.

While ultra-widefield imaging of the temporal retinal vasculature is most consistently captured with the ‘flying baby’ imaging technique, we also evaluated the advancement of nasal vascular front where data was available as part of an initial exploratory study and found this to strongly correlate with advancement of temporal vascular front (Supplemental figure 1). On this basis, TVR was chosen as a clinically convenient surrogate marker for overall retinal vascularisation. Our findings demonstrate significantly (29%) slower rate of retinal vascularisation in eyes that develop type 1 ROP (Group C) compared to eyes with no ROP or type 2 ROP (Group AB). This is consistent with previous reports evaluating the rate of retinal vascularisation based on indirect ophthalmoscopy (Solans Pérez de Larraya et al. 2018), timing of vascular front reaching Zone II (Jang and Kim 2020) and contact wide-field imaging using RetCam (Padhi et al 2022). The major advantage of using UWF imaging is that it allows rapid, non-contact, accurate and longitudinal monitoring of retinal vascularisation from birth to mid/late infancy (up to the age of 50 weeks in our cohort), consistently capturing the temporal vascular front up to anterior Zone II (without the need to collage images).

The risk factors contributing to delayed retinal vascularisation in premature infants, and therefore increased risk of severe ROP, include oxygen supplementation or fluctuation (Rivera et al. 2011; Hartnett and Lane 2013; Poppe et al. 2023), mechanical ventilation or CPAP (Allegaert, De Coen and Devlieger 2004; Yau et al 2016; Chan et al 2018; Solans Pérez de Larraya et al. 2019) and poor postnatal weight gain (Hellström et al. 2009). It has been proposed to result from a drop in serum insulin-like growth factor (IGF-1) level due to loss of maternal supply and poor endogenous production during extreme prematurity.

Since IGF-1 has a permissive role in VEGF signalling, low serum IGF-1 prevents retinal vessel growth in response to VEGF produced by the ischaemic avascular retina. As the infant grows bigger, endogenous IGF-1 production eventually catches up, thus permitting a rebound in VEGF signalling which drives the proliferative phase of ROP (Smith et al. 1999). Based on this model, postnatal weight gain has been proposed and shown as a biomarker for serum IGF-1 level and ROP risk (Hellström et al. 2009). Our UWF imaging-based method provides the most direct and consistent measurements of retinal vascularisation rate to date, which could be calculated from the earliest ROP screening visits (e.g. <32 weeks PMA) in extreme premature infants to facilitate early prediction and intervention to reduce the risk of developing type 1 ROP.

Our gradient boosting machine learning model, incorporating TVR along with BW and GA, was able to predict those eyes requiring ROP treatment with 96% balanced accuracy, 100% sensitivity, 92% specificity, negative predictive value of 80%, and positive predictive value of 100%, within a limited sample size validation test dataset. Whilst the prediction power of BW+GA and BW+GA+TVR models appear similar from the AUROC values 0.89 vs 0.90 (**Supplementary Fig.S4**), the more appropriate AUPRC values that consider the moderate class imbalance between non-treatment group and ROP requiring treatment group in our sample were 0.77 vs 0.84, which demonstrates an improvement following the addition of TVR (**Fig.3A**). Furthermore, the predictive power of BW+GA alone may be overestimated due to the limited sample size and particular nature of this cohort. A previous study obtained an AUROC for BW+GA of 0.808 (Chalam et al. 2011).

The addition of TVR to BW+GA for type 1 ROP prediction is also clinically meaningful as it improved the specificity of prediction from 87.5% with BW+GA alone to 92%, thus reducing false positive predictions. For instance, our combined TVR+BW+GA model correctly predicted infant ‘Test 9’ as not requiring treatment (Group AB), whereas the BW+GA model incorrectly predicted this infant as ROP requiring treatment, likely due to the infant’s BW and GA being below average for the validation set (803g and 26.4 weeks, respectively). Furthermore, infant ‘Test10’ is an example where, despite both BW and GA being higher than Test15, the combined model overrode this information and accurately assigned Group C based on low TVR (0.08 and 0.05) in both eyes. The variable inflation factor analysis also shows that TVR acts as an independent prediction factor over BW and GA (**Fig.3B**). Taken together, these findings suggest that TVR could provide added clinical value in ambiguous cases where it refines the typical correlation between BW/GA and ROP. Future work to identify clinical factors that modulate the rate of retinal vascularisation would also be beneficial for identifying novel therapeutic strategies to prevent ROP.

Across a cohort of 37 eyes (of 20 infants) that received intravitreal injection of bevacizumab (0.16 mg) for type 1 ROP between 32.5–39 weeks PMA, we found that anti-VEGF treatment does not significantly alter the rate of physiological retinal vascularisation. While there were individual examples of decreased TVR post anti-VEGF treatment, the overall trend of the cohort was in fact a slight but non-significant increase (Fig.4B). Sauer et al. recently reported that bevacizumab (0.25 mg) may increase vessel extension into peripheral avascular retina (Sauer et al. 2024). Therefore, both studies using similar methods of measuring the extent of vascularisation (from optic disc centre or optic disc edge, respectively) indicate that low or ultra-low dose intravitreal bevacizumab does not slow physiological retinal vascularisation.

Of note, the infants who did show reductions in TVR appear to be ones with unusually high pre-treatment vascularisation rates. Although there may be some lag in change of vascularisation rate after treatment, our follow-up period (mean 7.7 weeks ±7.9 SD) extended far beyond the half-life of intravitreal bevacizumab (shown to be 5-7 days) (Moisseiev et al 2014, Zhu et al 2008), including two infants observed for over 16 weeks without signs of reduced vascularisation rate. Future work is needed to identify what may determine the rate of temporal vascularisation and whether the heterogeneity in this characteristic may influence response to anti-VEGF treatment. Nevertheless, these intriguing findings challenge the assumption that anti-VEGF treatment slows physiological retinal vascularisation or causes vascular arrest, thus leading to the development of peripheral avascular retina (PAR) (Vural et al. 2019; Chiang et al. 2021; Hanif et al. 2022). This perceived disadvantage of anti-VEGF may encourage treating clinicians to delay intervention in eyes that are borderline for treatment (e.g. posterior Zone II, stage 2 with Plus or stage 3 with Pre-plus). Our findings provide novel insights on retinal vascularisation in premature infants with three main implications.

First, the results suggest that ultra-low-dose (0.16 mg) intravitreal bevacizumab may selectively target pathological extraretinal neovascularisation without significant interference of physiological intraretinal vascularisation. Second, the development of PAR is likely a result of slower rate of retinal vascularisation from birth in some extreme premature infants (as evidenced by our finding of slower TVR in eyes with more severe ROP), rather than a consequence of anti-VEGF therapy. PAR may have been previously under-recognised with laser photocoagulation to avascular peripheral retina but has become more apparent with increasing use of anti-VEGF as primary treatment modality. This has significant clinical implications for the aetiology of PAR and the timing of anti-VEGF treatment, suggesting the need to explore potential benefits of earlier intervention to suppress pathological neovascularisation (e.g. at stage 2 with popcorn lesions or in cases of retinal edge haemorrhage), which could prevent subsequent complications of more advanced ROP (e.g. fibrosis and vessel dragging that may complicate regressed stage 3 disease).

This is a single centre retrospective cohort study. Eyes with poor UWF image quality (that precluded accurate measurement of vascular front) were excluded, which could potentially introduce selection bias into the dataset. As a result, the gradient boosting predictive model was trained and tested on relatively small datasets. To further refine the prediction model, it would benefit from larger training datasets from multiple centres with prospective data collection. Our protocol for UWF imaging-based ROP screening has been successfully adopted at several neighbouring ROP screening units, which would help future improvement and validation of the model.

In summary, UWF imaging-based ROP screening enables continuous monitoring of retinal vascularisation rate, which provides an early biomarker for treatment-requiring ROP. The rate of retinal vascularisation does not appear to be affected by 0.16 mg intravitreal bevacizumab treatment, which has significant implications for the aetiology of peripheral avascular retina and optimal timing of anti-VEGF treatment for ROP. Future work will explore the effects of oxygenation and comorbidities on the rate of retinal vascularisation at different postmenstrual ages.

## Data Availability

All data produced in the present work are contained in the manuscript.

## Acknowledgements and Financial Disclosure

### c. Other Acknowledgements

Emer Chang and Amandeep Josan contributed equally as co-first authors.

## SUPPLEMENTARY MATERIAL

**Supplementary Table S1.** Raw data table of discrepancy between presumed fovea and actual fovea localisation using Flex OCT (as depicted in Figure 1). OD = right eye; OS = left eye.

| Patient | Eye | Discrepancy (mm) |
| --- | --- | --- |
| 1 | OD | 0.70 |
| 2 | OD | 0.29 |
| 2 | OS | 0.11 |
| 3 | OD | 0.30 |
| 4 | OD | 0.29 |
| 5 | OD | 0.20 |
| 5 | OS | 0.23 |
| 6 | OD | 0.00 |
| 7 | OD | 0.34 |
| 8 | OS | 0.40 |
| 9 | OD | 1.44 |
| 9 | OS | 0.50 |
| 10 | OD | 0.43 |
| Mean |  | 0.40 |
| SD |  | 0.36 |

**Supplementary Figure S1.**
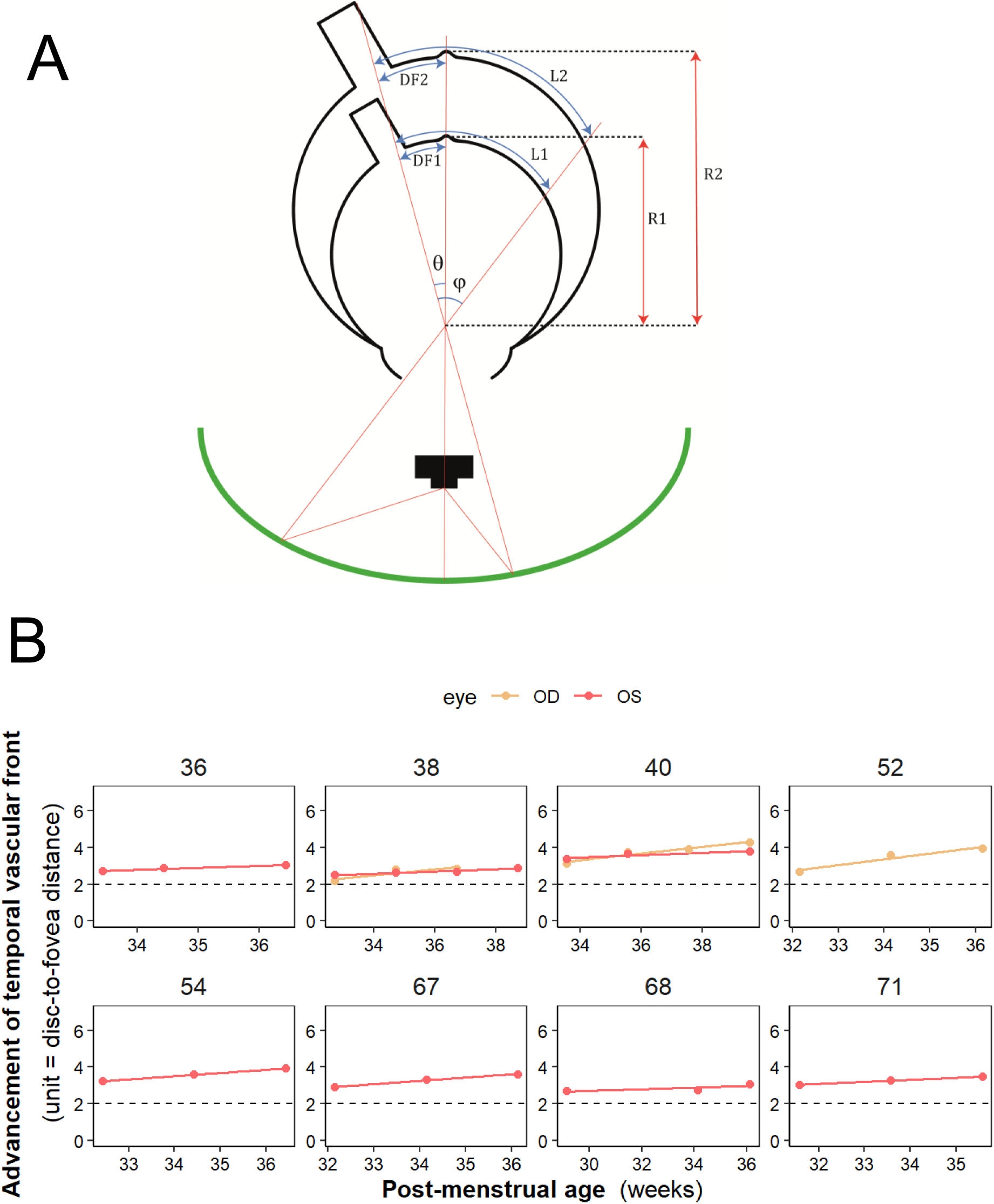
Ultra-widefield imaging optics and measurements. (**A**) Schematic of Optomap imaging of an eye at timepoint 1 (inner globe) and at timepoint 2 after axial length growth (outer globe). In a hypothetical scenario where the temporal vessel does not advance between timepoint 1 and 2 but only grows along with the eye growth/axial length, it can be shown that while temporal vessel length L2 is larger than L1 in absolute terms, the temporal vessel length expressed as a ratio to disc-to-fovea distance (DF2 and DF1) remains identical since DF grows in proportion to the vessel length. (**B**) UWF imaging-derived temporal vascularisation trajectory in untreated eyes of premature infants follow linear regression lines over the course of ROP screening (all available data shown for 10 eyes of 8 infants). Dotted line at y=2, where the ratio of disc-to-temporal vascular front distance to disc-to-fovea distance is 2, corresponds to the dividing line between Zone I and Zone II.

**Supplementary Figure S2.**
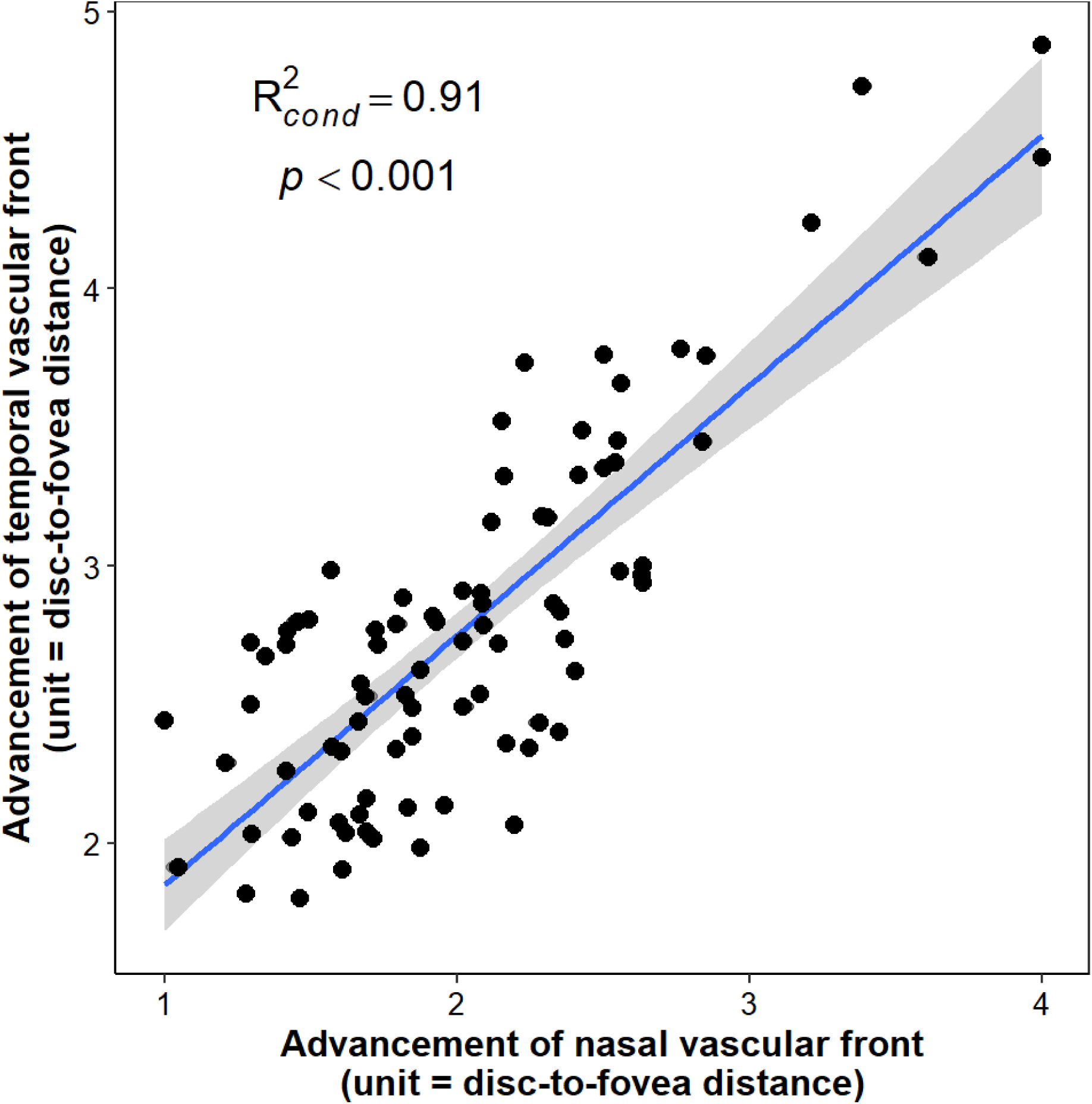
Significant correlation between advancement of nasal vascular front and advancement of temporal vascular front (p<0.001). Advancement of vascular front was measured from centre of optic disc to respective vascular front in disc-to-fovea distance units. R2= correlation coefficient. Blue line is regression line. Grey shading is 95% confidence interval.

**Supplementary Figure S3.**
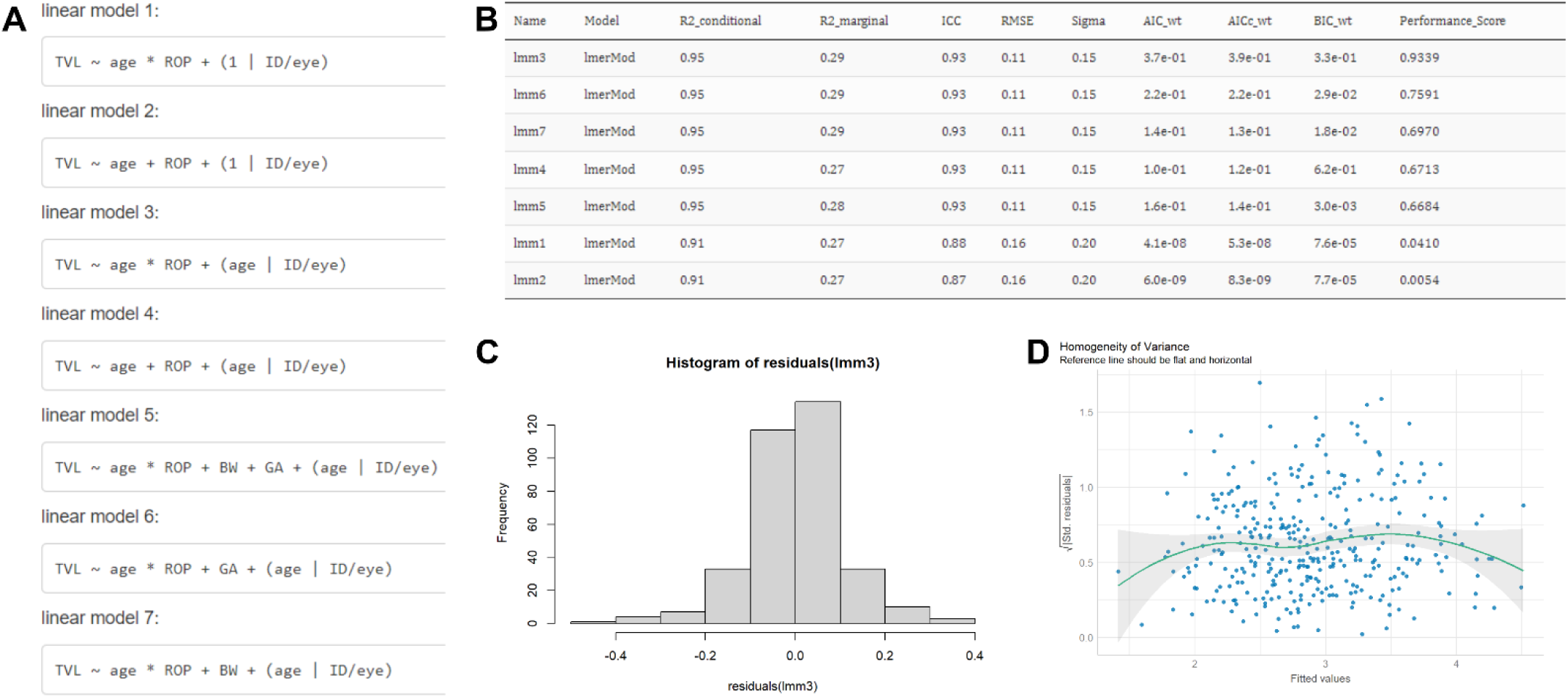
Model comparison and diagnostics. (**A**) Range of models investigated. (**B**) Model comparison table using Akaike information criterion (AIC) and Bayesian information criteria (BIC) to rank model performance. (**C**) Visual inspection of histogram demonstrating normal distribution of residuals of the final linear model 3 (lmm3), (**D**) Homogeneity of variance within confidence interval bands demonstrating no significant heteroscedasticity from lmm3. In addition to this manual model selection process, an automated brute force method investigating all possible combinations of predictors and their interactions also found linear model 3 to be the highest ranked model.

**Supplementary Figure S4.**
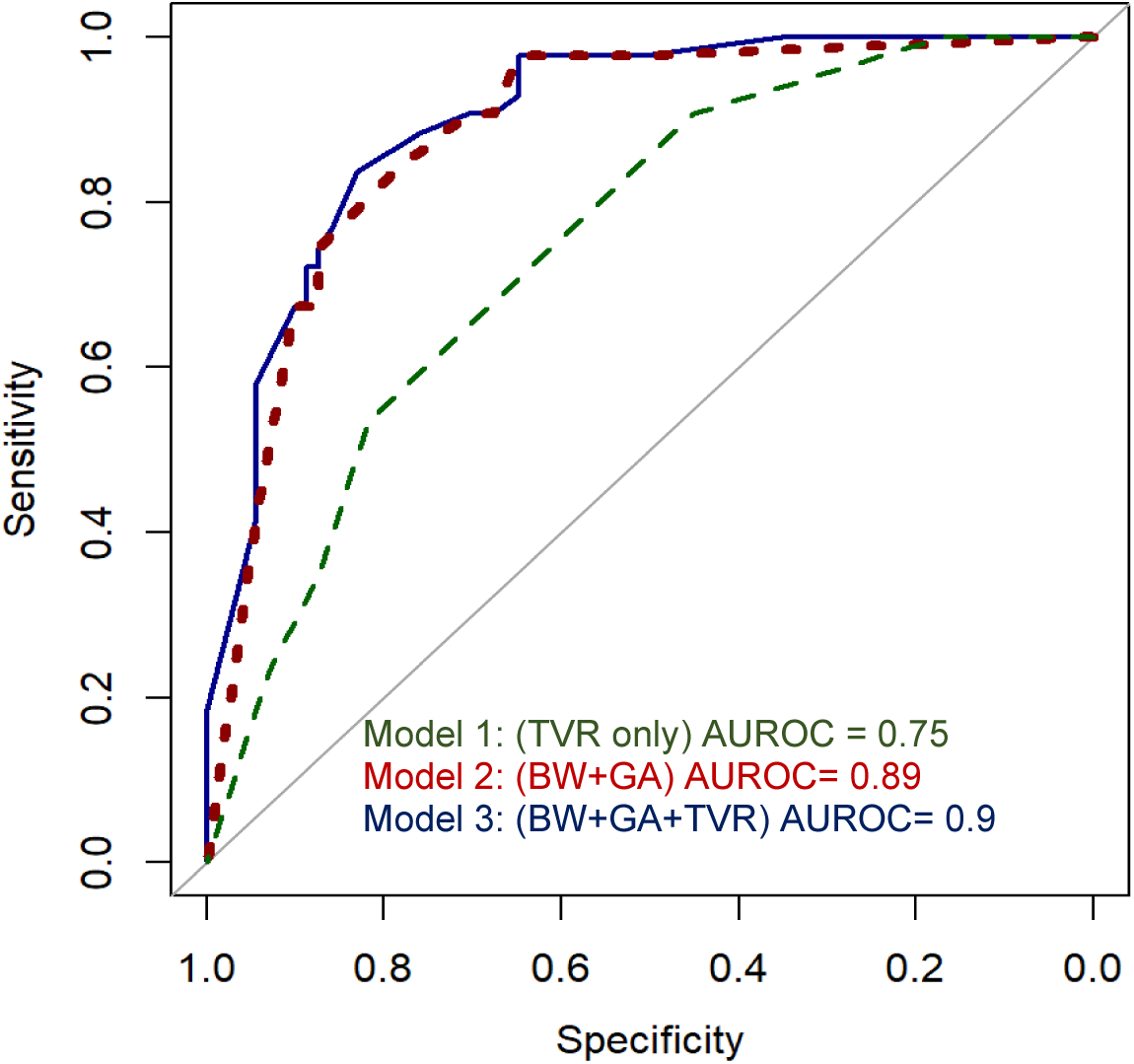
Comparison of the receiver operating curves for 3 different prediction models. AUROC: area under receiver operating curve. Dark blue solid line represents model with predictors as birthweight (BW), gestational age (GA) and temporal vascularisation rate (TVR) that was used for further prediction.

